# Roles, Challenges, and Solutions for Maternal and Child Health at rural Communities in Bangladesh

**DOI:** 10.64898/2026.10.18.26365859

**Authors:** Abu Sayeed Abdullah, Abdul Halim, Farjana Haq, Abu Sadat Mohammad Sayem, Koustuv Dalal

**Affiliations:** Centre for Injury Prevention and Research, Bangladesh (CIPRB), Dhaka, Bangladesh and Department of Public Health Science, Mid Sweden University, Sundsvall, Sweden; Independent consultant, Dhaka, Bangladesh; Independent consultant, Global Health Nexus, LLC; Department of Public Health Science, Mid Sweden University, Sundsvall, Sweden

**Keywords:** Roles, Challenges, Solutions, Maternal Health, Child Health, Bangladesh

## Abstract

The rural community has an important role in improving the maternal health and child health situation in Bangladesh.

A research study was conducted in 2020 to explore the impact of Community people on maternal and neonatal health seeking behavior at Kalaroa upazila of Sathkhira district in Bangladesh. A comprehensive study involving two focus group discussions (FGDs) and sixteen in-depth interviews (IDIs) was conducted with Community Group (CG) and Community Support Group (CSG) members to delve into their experiences, duties, and challenges as members of their respective communities.

The participants highlighted their responsibilities, organizing meetings, providing health advice, and community support. They also discussed encountering obstacles, such as community skepticism towards family planning and financial inquiries, particularly from affluent individuals. Overcoming these barriers involved proactive communication, collaboration with higher authorities, and continuous community engagement. In terms of enhancing maternal and child health, community emphasized the need for improved healthcare infrastructure, including access to qualified medical professionals, essential drugs, transportation facilities, and better-equipped delivery rooms at community clinics. Some also advocated for separate funding mechanisms and enhanced training for staff.

Overall, their insights underscored the crucial role of community in promoting health addressing challenges to achieve better outcomes for mothers and children.

## Introduction

Bangladesh has experienced a steady decline in maternal and under-5 child mortality during the recent decades representing progress towards achieving the sustainable development goal (SDG)^1^. The maternal mortality ratio (MMR) fell from 322 to 136/100,000 live births between 2001 and 2023 ^2^ while neonatal mortality declined from 52 to 20 deaths per 1000 live births^3^.

Reproductive health has been identified as an area of high priority in the Health, Population and Nutrition Sector Development Programme (HPNSDP 2011-2016)^4^. The operational plan of maternal, neonatal and child health has identified the priority areas, that includes improvement of the quality of EmOC services from all levels of hospitals, social and community mobilization, revitalization of Community Clinics (CC) and improved participation of community for CC management through establishment of community groups^5^.

Bangladesh has achieved considerable success in the reduction of maternal and neonatal deaths over the past decade. Though the reduction of maternal deaths has been the focus of most interventions, the neonatal mortality rate (NMR) also remains considerably higher than expected. The current NMR is 26 for every 1,000 live births and the target has been set to 18 per 1,000 live births to be achieved by 2022 and to 12 per 1,000 live births by 2030, under the Sustainable Development Goal (SDG) 3.2^6^.

Bangladesh has made significant strides toward social transformation and poverty reduction and is now expected to leave the category of least developed countries within the next five years. Social investments made to promote adolescent girls’ education and give women more clout, is mostly responsible for the advancement.^7^

Between 1990 and 2013, under-five mortality decreased by 72%, while neonatal mortality decreased by 55% in Bangladesh^1^, Stronger efforts are required to bend the curve to achieve national NMR and still births target of 10 or less per 1,000 live births by 2035. There is also a progress in postnatal care with 36% of mothers and children receiving check-ups from a medically trained provider within 42 days of delivery.

The national sector programme: HNPSP and the current HPNSDP 2011-16 raised the effective coverage (quality) issues as priority gap to resolve immediately particularly in maternal and neonatal health. In the recent years, Government of Bangladesh (GoB) has been implementing in partnership with UN organizations and development partners a few innovative initiatives at district level which created enthusiasm in making progress. Joint GoB collaborative MNH Initiatives (MNHI), MNCS, and MNCH programmes. The joint efforts in MNCH created an enabling environment and mobilize ample resources to encourage innovations, develop of human resource, strengthen referral system, timely ensure supply-procurement of logistics and drugs etc. for better performance. Quality improvement is central to progress in MNH services.

Adolescent females experience significant assault and exploitation due to damaging cultural standards.^8^ Whereas when it comes to adolescent education in Bangladesh, it depicts different picture. Adolescent girls enter in secondary school at a larger rate than boys do (74 percent vs 62 percent, respectively), most likely because of the government’s tuition assistance and stipend programs for female students, but their dropout rates are frightening. Nevertheless, only 68 percent of adolescents were engaged in secondary school on a net basis in 2019. The same study reveals also geographical disparity alongside gender inequality. ^9^

The overall objective of the research was to understand the Roles, Challenges, and Solutions for maternal and child Health at rural Communities in Bangladesh. To comprehensively investigate the landscape of maternal and child health initiatives in rural communities of Bangladesh, with a focus on stakeholders, challenges, service accessibility, intervention effectiveness, and the development of actionable recommendations for system strengthening.

## Methods

A qualitative study was conducted in Kularoa Upazila of Sathkhira district of Bangladesh during October 2020 where focus group discussions and in-depth interviews were performed^10^. A total eleven focus group discussion (FGD) and sixteen in-depth interviews (IDI) were conducted within the period of time. For FGDs, we chose five groups. First group was with the mothers of intervention areas where six FGD were performed; second one with the mothers of control area where two FGD were performed. The selected mothers for FGD of intervention and control area were those who delivered a live baby during the period of 1^st^ July 2019 to 30^th^ June 2020. The third and fourth groups were selected from the member of community support group and community group of community clinic in intervention areas. In the last group was with the community volunteer of that intervention area. Each of group consist of 9-11 participants. Sixteen IDIs were performed with the member of community support group and community group of community clinic in intervention areas with a few ten and six respectively. All IDIs were conducted following guidelines by face-to-face interview at the household level. The tools and guidelines were drafted through small group discussions with the experts in CIPRB under the guidance of the PI and Co-PI of the study.

Two trained research officers including an anthropologist were assigned to collect the data from the field. Formal training was provided; guidelines were pre-tested before data collection. During FGDs, one research officer facilitating the discussion whereas, other researcher took important notes. The objectives of the research were clearly demonstrated to respondents before the interviews. A written consent was taken from each of the respondents before the interviews or FGDs. A few prompts were used to obtain the information. Audio voice recording was done prior to permission from the respondents. From the audio-recordings and hand notes of the interviewer’s, the research officers prepared IDIs and FGDs transcripts in native Bengali language. Peer debriefing was also performed to maintain its reliability of the data. Initial open code was done, then from those open code, selective coding was done. Themes were identified after reading and re-read of the data and finally analysis was performed thematically.

**Table 1.**
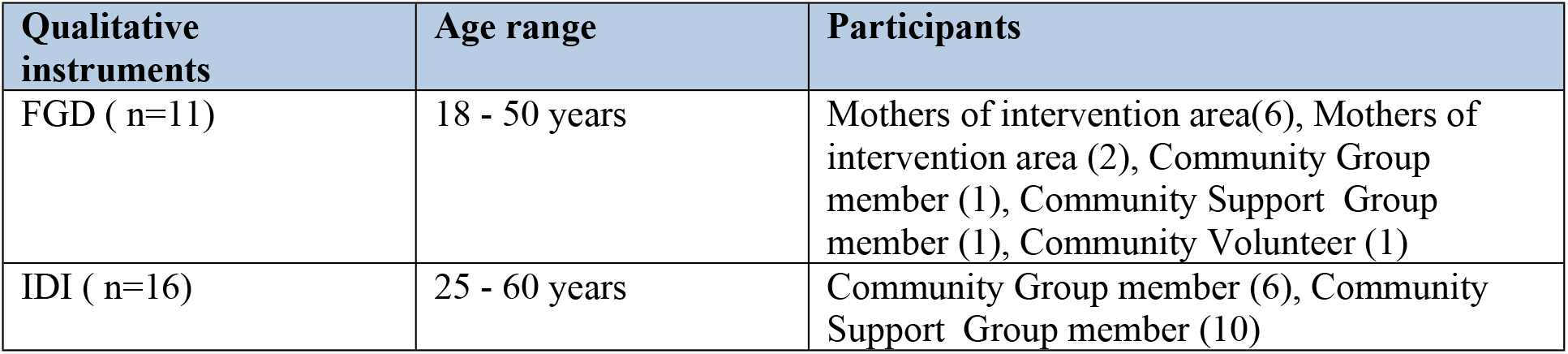
List of Participants in the qualitative study.

| Qualitative instruments | Age range | Participants |
| --- | --- | --- |
| FGD ( n=11) | 18 - 50 years | Mothers of intervention area(6), Mothers of intervention area (2), Community Group member (1), Community Support Group member (1), Community Volunteer (1) |
| IDI ( n=16) | 25 - 60 years | Community Group member (6), Community Support Group member (10) |

**Table 2.** Content of the focus-group discussion and In-depth interview.

| Group | Area of discussion | Types of Prompts used |
| --- | --- | --- |
| Mother s in the Intervention and control area | Experiences of last pregnancy | Feelings of last pregnancy? Idea and practice of birth planning? Idea and experience of antenatal checkup? Problem faced while taking services? |
|  | Experiences of last delivery | Where delivered and how? Who assist delivery? Any problem during delivery or while getting services? |
|  | Experiences in post delivery | Feeling and practices after delivery? Problem of responders or her baby? Why the responder didn't receive PNC? |
|  | Knowledge about CSG member and Community Volunteer | Idea about CSG & Community Volunteer? Ideas about the activities of CSG & Community Volunteer at community? |
| CG and CSG members in the Intervention area | Idea and practices about their responsibility | Feeling to be a member of CG/ CSG? Responsibility of CG/ CSG? Practice to fulfill these responsibilities? |
|  | Barrier on the duties as CG/ CSG and practices to Overcome | What are the social and family barriers at community to fulfill the responsibilities of CG/ CSG? Practices to overcome the barriers? What else they can do? |
| Community Volunteer in the Intervention area | Experiences of a community volunteer | Reason to be a community volunteer? How did they help mother and children as a volunteer? |
|  | Barrier on the duties as community volunteer and practices to Overcome | What are the social and family barriers at community to fulfill the responsibilities of community volunteer? Practices to overcome the barriers? What else can be done for the improvement of mother's and child's health? |

The documents were prepared for submission to the CIPRB ethical review committee (ERC). These documents included the Umbrella Proposal-Initially submit an overview., Protocol, Abstract Summary for Ethical Review Committee as per the attachment, Informed consent form for subjects, informed consent form for parent or guardian, Written Consent form for subjects, Procedure for Maintaining confidentiality, and Questionnaire or interview schedule. The proposal was presented in the ERC committee meeting with the support of the PI and CO-PI of the study. The feedback of the ERC board members was incorporated into the proposal and resubmitted to the ERC.

A quality control team consisting of senior research team members from CIPRB were deployed for quality control checking of the study data. Quality control checking were designed to physically verify whether the data collectors completed the interview and FGDs by interviewing the right respondents by asking the right questions. Quality control checking were undertaken both in ‘presence’ and ‘absence’ of the interviewing team. After data collection, all the qualitative findings was processed under supervision of research coordinator & team leader.

All the qualitative findings were processed under supervision of team leader. Audio recordings were transcribed independently by two anthropologists and compared for accuracy. Transcriptions were coded independently by the two research assistants. Transcription were made in Bangla and subsequently were translated into English. Using content analysis major themes were identified and coded. After sorting and categorizing the responses, excerpts from the transcripts were chosen to illustrate the summary statements, which were used to validate the findings. As stipulated in the ToR, the research team assisted in the preparation of the report under the guidance of PI and Co-PI.

## Results

### MNH services in mothers’ group: FGD – intervention and control

Eight Focus group (FGD) discussions including six FGDs with mother of intervention areas and two FGDs with mothers of control areas was conducted at Kularoa upazila of Sathkhira district. These FGD were conducted to compare the knowledge and practice on maternal and neonatal health related services between the mothers of intervention area with those of control area. These FGDs revealed higher level of knowledge and practices on birth planning, expression of feelings & complication during pregnancy, receiving antenatal and post-natal care during pregnancy among the mothers of intervention area. Whereas the mothers of control area had misperception on birth planning and had no ideas on maternal complication during pregnancy.

#### Feeling to be a pregnant mother

In response of the feelings during pregnancy most of the mothers of intervention area clearly expressed their experience during pregnancy. They can detect earlier if any complication arises during pregnancy and receive care. Whereas most of the mothers of control area cannot recognize pregnancy complication as they thing these are the normal condition during pregnancy. They feel scared but happy to be a mother during pregnancy period.

One of the mother of control group said *“I couldn’t eat anything even medicine provided after checkup. Sometime I might eat two days interval as I always had vomiting tendency and my legs were swelling with fluid. I felt afraid during pregnancy either my baby might born healthy or born with any problem. Our village doctor said that these are not problem. But when I felt about to be a mother then feels good. During delivery there was problem as the traditional birth attendant could not remove the placenta. Then I was brought to Kesobpur Private Clinic where I need two bags blood*.*”*

During FGD one mother of intervention area said *“I felt good for going to be a mother for the first time. All the member of my family was happy for that. I may not receive education but always dream to make educated of my baby. When I felt any complication then I discussed with community volunteer. I suffered from physical weakness & headache during my last pregnancy. Then I went to community clinic for antenatal checkup. When ultra-sonogram report showed that my baby was in good condition then I felt happy*.*”*

#### Birth planning issue

On the responses of birth planning for delivery, mothers from the intervention area explain their better reaction. They select the place & person for delivery previously, receive ANC checkup regularly during pregnancy, communicate with community volunteer and preparing vehicle. On the other hand most of the mothers from the control area have no importance on birth planning. Although few of them responses their contact with the local tradition birth attended for home delivery.

One of the mothers of intervention area said *“I received all four ANC from community clinic. Moreover I had a health card mentioning the preparation for delivery like arranging emergency vehicle facility, saving of money etc. I followed these like blood test, ultrasound test and weight measure regularly. I saved money & went to my father’s home near to the Kularoa hospital before delivery*.*”*

One of the mothers from control area said *“we contacted with the local tradition birth attendant for delivery at home during my last stage of pregnancy. If any problem we might go to hospital. But we had no saved money. We didn’t think to ready vehicle earlier*.*”*

Another mother from the intervention area said *“I had excessive water in my body, so I cannot walk though I was check up monthly in the community clinics and I had blood tests and other tests before delivery, I had also done ultrasound for seizure in the clinics. I had a wish to delivery at home by the skilled birth attended. A vehicle was ready for emergency transport. Moreover I had mobile no of MBBS doctors to contact. I received ANC regularly from community clinic. My blood group & ultra sonogram test was done at last stage of pregnancy*.*”*

#### Knowledge and practice on antenatal care

In response to the information about the knowledge and practices on antenatal checkup most of the mother of intervention area received ANC regularly including health checkup, blood test, ultra sonogram. On the other hand the mothers of control area had no clear idea and importance about antenatal checkup. They believe and practice to go at hospital if any complication arises.

During FGD a mother from intervention area said *“I received four times ANC during my last pregnancy on third, sixth, seventh and ninth months of pregnancy. The doctor checks my blood pressure, body weight & movement of my baby during ANC. I avoid doing any hard work and taking rest as doctor advice me. I always everyone in house was helpful*.*”*

One of the mothers from control area said that *“I didn’t go to hospital as no problem arises. At first I received ANC on ninth month of pregnancy though I couldn’t detect the fetal movement*.*”*

#### Complication of pregnancy

Most of the mothers from intervention area expressed their complication during last pregnancy easily mentioning abdominal pain, headache, weakness, problem to walk and daily work and edema in the body. Mothers from the control area mentioned huge pain and high pressure problem and problem occur during delivery. But they things these are normal phenomena during delivery.

One mother of intervention area said *“I had severe headache during last pregnancy and my feet & face was swollen. Doctor advised for caesarian section & I agreed to do*.*”*

One mother of control area said *“My baby was injured in her head due to force delivery at home by unskilled birth attendant. My legs were swollen during pregnancy*.*”*

#### Obstacles to receive antenatal care

Most of the mothers from intervention area do not face any obstacle during pregnancy. Their family members know about the importance of receiving ANC. But the mothers of control area faced lots of obstacle during ANC both from family and society.

One mothers from intervention area said *“Now we are aware to receive ANC. Earlier mother in laws prohibited receiving ANC but now they insist us for ANC*.*”*

One mothers from the control area said *“mother in law is the main barrier to follow the advice of doctor. She said about the difficulties of delivery of large baby for eating more during pregnancy*.*”*

#### Feeling during delivery

The mothers from intervention area express their happy feelings of being a mother during delivery. The mothers of control area mention various difficulties situation during delivery.

One mothers from the intervention area said *“Two begs blood need during my delivery but when I saw my baby I forget my all pain*.*”*

One mothers from the control area said “There was bleeding for placenta tear after delivery at home by traditional birth attendant. Then I was carried to hospital where doctor said about my life thread”

#### Time and place of delivery

Most of the mothers from the intervention delivered safely at private clinics or government hospital. Few of them said their delivery took place at home with skilled birth attended. However most of the mothers of control area delivered at home by traditional birth attended or village doctor.

One mothers from the intervention area said “I delivered safely at Kularoa Clinic by caesarian section. I plan this previously with the discussion of my family members.”

One mothers from the control area said “I delivered at home by TBA but after delivery when bleeding arises then I was carried to the Kularoa hospital.”

#### Obstacle faced during delivery

The mothers of intervention area have no problem during delivery as they were aware previously. However the mothers of control group faced lots of problem during delivery as they delivered by traditional birth attendant at home. Maximum of intervention areas mothers delivered safely at facility.

One mother of the intervention area said *“I face no problem during delivery at clinics. I was aware about problem during delivery*.*”*

One mother of the control area said *“I delivered 24 hours after pain. At first my mother in law tried to assist. When complexity arises then they called the village doctor*.*”*

#### How the mother take care the neonates

Most of the mothers of intervention area are careful about Childs hygiene, allow to bath after three days of delivery, provide colostrums milk immediately after delivery. These mothers have the ideas on neonatal care from community volunteer. They can detect earlier if any complication arises in neonates. Mother from the control area allow bath to child immediately after delivery and also give honey to eat after birth.

One mother of intervention area said *“I give breast milk to my baby immediately after birth. I allow my baby to bath three days of age. I know six danger sign of neonates from community volunteer*.*”*

One of the mother of intervention area said *“my baby suffer from fever after three days of birth then I took him at hospital where infection was detected in blood and injection required for his treatment*.*”*

During FGD one mother of control area said *“my baby suffered from pneumonia after birth. At first coughing and fever occur after 4-5 days of age but we can’t understand*.*”*

One mother of control area said *“my mother in law allow to make bath of my baby immediately after delivery to clean its body. I was so weak but I was so happy after see my Childs face and forget my all pain and sufferings*.*”*

During FGD one mother of intervention area said *“after birth I cover my child with clothes for make hot, as I had no brest milk so gave powder milk by the suggestion of doctor. After birth I cleaned my child napkin and dry. I also give him colostrums milk immediately after birth for immunity. “*

#### Postnatal care

Most of the mother of intervention area received postnatal care immediately after delivery as they knew this earlier. On the other hand most of the control mothers think its need money for postnatal care.

One mother from intervention area said *“I delivered at hospital where they provide PNC and after 15 days I came again to receive PNC again*.*”*

One mother from control area said *“I delivered at clinic and after eight day back to home and never went there for any further checkup. We had no enough money for postnatal checkup*.*”*

#### Idea about the Community Support Group and Community volunteer

In response to the knowledge about the Community Support Group and volunteer the mother of intervention areas responses well. They always have communication with volunteer as they organize the court yard meetings for awareness on maternal & child health. About their activities at community most of them mother from the intervention areas said about arranging courtyard meeting, advising pregnant mother about feeding, hygiene and check up during pregnancy. But the mothers of control areas have no ideas about the activities of CG, CSG and volunteer. Some of them also said that they never hear this person from anywhere.

During FGD one mother of the control area said *“We don’t know about CG, CSG and we have no communication with them. No one inform us about them*.*”*

One mother of the intervention area said *“they made yard meeting, said how to eat in pregnancy, how to wash hand, how to feed children and washing hand with soap. They told us to take rest, tell about sleep 8 hourly. Once my neonates had convulsion and Amedi apa, volunteer, arranged a motorcycle for transport by150 taka. Sometimes she go with us to the clinics and instructed us where and how to go*.*”*

One mother of the control said *“there was no yard meeting, no meeting mother, no one can get money during operation and there is no DSF/yellow card in our area. During pregnancy I participated in a meeting near to mosque but who did this I don’t know*.*”*

During FGD one mothers of the intervention area said *“Rojina and Fulmoni are the volunteer in this areas. They arrange the courtyard meeting where they aware us about the maternal and neonatal care, where to deliver the neonates, how to take care of the neonates. They also show us the pictures of maternal and neonatal danger signs to aware us. Mutalib Morol is the member of main committee of our community clinic. We don’t know anything more about CG and CSG*.*”*

### FGD findings from community volunteer

Two FGD were conducted with the community volunteer of intervention area to understand the reason to be a volunteer, ways of support mother & children at community, barrier to work as community volunteer & overcome method & suggestion to improve the mother & child health at community. They feel proud to be volunteer and satisfy to serve for the betterment of mother & neonatal health. Though face some family & social obstacle but can overcome with their continuous services. They suggest improving the facility of community clinic for the betterment of mothers & child.

#### Reason to be a Community Volunteer

In response to the opinion about the reason to be volunteer most of them mention about working for the welfare of mothers and child in our areas. They also mentioned this is the opportunity to benefit of the mothers. They work as volunteer with their own interest without any outside pressure as everybody appreciate for a good deal. They can learn new things and disseminate the knowledge among the community.

During FGD one volunteer said *“many mothers and child died here earlier. But now we get the opportunity to save their life & no more mothers and child will die here. I heard first from a sister of JICA office to work here for the welfare of mothers and Childs. When she asked about our opinion to be a volunteer about this program I feel very interest to involve here*.*”*

During FGD another volunteer said *“there are lots of illiterate people in rural community. We have plenty of time beside our regular work. In the mean time we can provide services to the community people. Pregnant mothers didn’t know many things due to their illiteracy. We can disseminate the knowledge among the village people after receiving training from the office. If we spend our lesser time for mother’s welfare then they can be benefited and that’s why we work as volunteer*.*”*

#### How community volunteer help mothers and children

In response to the contribution of helping mothers and child many of them mentioned about the receiving advice on antenatal checkup, inquiring us for advice during complication of mothers and child. The rural people are motivated to receive the health care from community clinic and Upazila health complex. The volunteer distributed the card among the mothers where golden card for make them understand about the ANC care, safe delivery & birth planning, yellow card contain five danger sign of mothers and green card contain six danger sign of child and also help in quick referral of high risk mothers.

During FGD another volunteer said *“we didn’t know earlier about the role of a volunteer. But when Suvas sir said if you are interested to be a volunteer then it is easy to disseminate the knowledge among the community to save the lives of mothers and child. Though we may face lots of problem but when we thought we can save the life of mother through volunteerism then we were interested to be a volunteer. Being a volunteer we can learn a new thing and teach others also about that thing. Even it is important for our own life as this knowledge is useful for saving the life of our own mothers, sisters, brother in laws*.*”*

During FGD another volunteer said *“we didn’t find interest at first as the people ignore us even said that these things are not necessary for us and didn’t want to provide their name for registration. They also searched some financial support from us. But now people search us with their own interest. If someone became pregnant then call me for advice & requested me to register their name. Now hues participation occurs in the courtyard meeting. When any complication arises at home delivery then we obtain the nurses from hospital*.*”*

During FGD another Volunteer said*” maximum people have no capacity to transport by ambulance then we manage the local van, rickshaw for immediate transport. We sometimes also go with them and stay at hospital with them. Earlier many mother died after delivery of baby which is due to ignorance of mother care after delivery. Now we make them understand about the importance of caring mother immediately after delivery by showing them the cards containing the danger sign of mothers. The mother didn’t know about the danger sign of neonates. We show them the danger sign of neonates. Now they can detect either the baby is healthy or not and take the hospital if any neonatal complication arises*.*”*

During FGD one Volunteer said *“we received training on maternal and child care. We learn practically the process of neonatal care immediate after delivery with the help of a dummy doll. We learnt how to clean and warm up the neonates immediately after delivery. Then we demonstrate these in front of the villagers & they now practiced it. Mothers didn’t know about how many times need to receive the ANC checkup and from whom and where but now they can understand. We continuously said them to receive ANC from community clinic or Kulaora UHC. Now they receive our advice and go at community clinic for receiving services and medicine. Sometime they fall in trouble for emergency transport of patient as they had no number for calling Ambulance, then we call ambulance for taking the patient immediately to the facility*.*”*

During FGD another Volunteer said *“We took the information of last date of menstruation and mark in our notebook. At first we registered the mother immediately after pregnancy and visit four times in a year. We advice them about movement during pregnancy, food intakes, vaccination, taking rest and avoid hard work. Advice them for the preparation of delivery like collecting sterilized blade, cloths, savlon and also save little amount of money regularly. We also advice them about the PNC care within 24 hours after delivery and avoid bath of neonates within 72 hours. Provide colostrums to neonates within 01 hour*.*”*

#### Challenges and obstacles to be a volunteer

In response to the challenges and obstacles they face during volunteerism they said that the community people didn’t give important on their advice even some times ignore them. The community people didn’t easily break their traditional practices. They expected some financial support when the volunteer visited their home.

During FGD one Volunteer said *“the mother in law occurs the main problem in the family & said that they understand all the things without our help. Even the mother didn’t want to say the last date of menstruation as they thought may occur problem in their pregnancy*.*”*

Another one said *“At first we got obstacles from our own home. Our family member asked about our objectives of going at door to door and always discourages us to participate. The neighbor said what benefit and money you received for doing this work. They can’t believe that we provide services without any benefit*.*”*

Another one said *“sometimes mother in law said I delivered three to four times without any problem. We practiced hard work during pregnancy but now you said not to do any hard work. BRAC provide us plate, pot, money and feed but what you will provide us. Why you come without any things?”*

#### How to overcome the obstacles

In response to overcome the problem and challenges many of them said that they need to cool in term of the neglected behaviors of the responders. At first the community people didn’t want to learn anything’s from us then they need to go there several times to make them understand about the importance of this program. They need to visit door to door for sensitization of community. Many pregnant mothers veil their concept then they need to counseling with them. Gradually when the community people benefited then they show their faith on them. They also inform the CG members for their help in solving the problem. Even they involve the elite person of the society to aware about receiving the vaccine of mothers & child.

During FGD one Volunteer said *“At first we became fail to make the people understand about our activities. But we didn’t stop there. If we didn’t move, then the work will become stop. Then we visit door to door for two to three times or even five times in some houses with a hope that once they will provide importance on our activities*.*”*

During FGD another one said *“we face many obstacles earlier then we shared these in bimonthly meetings. We requested the community support group member to facilitate the male guardian of pregnant mothers at bazar or where meet. Then they motivate their wives not to ignore our advices. Earlier the guardian became angry to hear about the vaccine even said us to go out from their home as they believed that the death occur due to vaccine. But now they are aware about receiving the vaccine. Even we requested the Imam of mosque to say their speech for providing vaccination of their kids*.*”*

During FGD another one said *“We make them understand with patience. Sometime they became rush on us but we make us cool. We said them we come here only for their well beings. But when someone benefitted from our advice then they become respectful on us. Mothers hide their pregnancy up to 3 to 4 months. Then we need to counsel them about the antenatal cares and maternal complication. At 9 months of pregnancy they thought that we went there to provide them necessary equipments like blade, soap etc. Then we explained our work and provide advice for safe delivery & postnatal care*.*”*

#### How to improve the maternal and neonatal health

In the response of what else can be do for further improvement of health of mothers and child may of them said that the community clinic need to open for 24 hours for providing services with availability of essential drugs and oxygen. There is necessary of expert employee including doctors should appointed at community clinic. Some of them also said it needs to expand the areas of community clinic. They also emphasis about to provide their honorarium for making them more involve here.

During FGD one volunteer said *“it needs a better place for delivery at community clinic. The bed should be provided for patient. Toilet & tube well need to construct. Electricity, water supply is essential here. Here it needs a gynae expert for delivery or caesarian section at community clinic. The quantities of drugs also need to increase, even oxygen supply need to ensure. “*

During FGD another volunteer said *“we are working here for more than two years as volunteer only with our personal interest. But if we got some honorarium then we will encourage for more involve here with responsibility. We can learn more if monthly meeting & refresher training are organized*.*”*

During FGD another volunteer said *“A vehicle should provide for transportation of risky mother. Sometimes mother may feel delivery pain for a long time but can’t reach to hospital for scarcity of vehicle. The mothers are not normally interested to go at facility at far away from home. If transport facility is available here then our patient can easily go at clinic or hospitals*.*”*

### FGD and IDI outcomes from CG/CSG members

A total of two FGD and sixteen IDI were conducted with the CG & CSG members to understand their feeling to be a CG/CSG member in the community. The duties & responsibilities of CG/CSG in community also can be recognized. They also share the obstacle faced at community & how to overcome & provide suggestion for improvement of mother & child at community. One FGD was conducted with CG members and other one was conducted with the CSG member. Among these sixteen IDI six were conducted with the CG member & ten were with CSG members. During FGD and IDI many of the participants expressed their better feelings to be a CG/CSG. They also mentioned maximum responsibilities of CG/CSG and their specific duties at community. Maximum of them mentioned the traditional ideas are the main social barrier and continuous motivation is the main way to overcome these obstacles.

#### Feeling to be a CG/CSG member

Most of the CG & CSG members explained their better feeling as people respect them. Though the community people cannot understand anything at first but now they can appreciate everything. They also feel good to advice the mothers & helping them during their critical condition. Some of them feel proud to be a CG/CSG member.

During FGD one CG member said *“It is very charming work that feel us very happy. In the health sector government made a huge improvement and by joining here I am very pleased. I like it but only due to mothers, family and babies are benefited. Last year one pregnant woman was in difficulties and I helped her by renting a car and send her to the Kularoa hospital for health facilities and the peoples of this area are also known me as a good man*.*”*

During IDI one CSG member said “*I liked to help people. I stand beside when they are in trouble. Previously mother and Childs died but now days they are not. If they take my name then them given drug, they are getting so much happy when they show their house to house. There are many people in society who are more rich & educated than me but they want suggestion from me. I like to help peoples of my own places, such as one day a pregnant woman had delivery pain but she stayed at home and I convince her to delivery at health care centre. “*

During FGD one CSG member said “*I feel so exited, so exited and there is no bound for happiness to help others. We advice the community people not to give honey to baby’s mouth, not to hold baby with dirty clothes, not to bath babies immediately after birth. I have to talk a lot with them; they are unable to understand now they can realize the importance of our suggestion*.*”*

During IDI one CG member explain *“When I helped people in their difficult periods then it makes me happy, the people of this area get helped by taking drug from this community clinic. When we go to village then peoples are explain their facilities and problem and we are suggest them to come to clinics. I always try to overcome people from their sin and if there any mother get sick then I advice her to go to Kulaora hospital. I was trying a lot for a mother to take her in the doctor but they didn’t go and at last baby was died. That time road was very rush and deficiency of vehicle. Now a day’s people have come to us by their selves because they are aware now*.*”*

#### Knowledge about the responsibilities of CG/CSG

In responses to the knowledge of duties and responsibilities of CSG and CG most of CG mentioned about conducting meetings regularly, solving problem in the meeting, and advice mother on health care. Some of them said about fund raising. CSG illustrate their responsibility as arranging meeting regularly, advice about danger sign among pregnant woman and take care of baby, to take mothers and child to the community clinic if necessary and make fund for serving patient’s critical condition.

During FGD one of CG member said *“We conduct monthly meeting, discussed problem in the meeting, keep record of the drugs, when someone take medicine then I talk with him on the road to see what drugs are given. We also participated in the meeting arranged by Ahsania Mission & Nari Pachar, advice to provide vaccine, searching the illiterate mother who forgets to receive vaccine regularly*.

During FGD one CGS member mentioned *“We advice the pregnant mothers about taking rest for a healthy baby, extra food intake, avoiding risky work. When babies are under nutrition then we advice them as if babies are less than 800 gm weight or navel ill then go to doctor. The danger sign of pregnant woman like excessive bleeding, fever and headache these cases are very sensitive and I suggest them to go at Batra community clinic. I referred the risky mother for safe delivery at facility*.*”*

During IDI one CSG member said *“we made a fund for savings at community. From the main comity we save monthly, if government left this clinic near future then can we run this? That’s why we take rule that we can run this by ourselves. That’s why we are made a big fund. We made a fund in the community clinic to help poor mother for caesarian operation. We advice to avoid hard work during pregnancy which is also harmful for her baby*.*”*

#### What they do as CG/CSG member at community

In response to how they fulfill their duties most of CG organized meeting for discussion many issues. Few of them investigate the problem and supervision of CGS members. The CSG members arrange meeting, advice the community people regarding the care of mothers and child.

During FGD one CG member said *“I attend meeting regularly. I discussed about community clinics with the other member of community clinics in the market. Sometime I gave my own money if there any emergency reference of patient*.*”*

During FGD one CSG member said “*We collect the community problem from community volunteer and organize a courtyard meeting to solve these problems. I advice the husband group to save money during emergency, prepare for transport and blood for emergency. I also discussed about five danger sign of mothers and six danger sign of child*.*”*

During IDI one CG member said *“I call every member for meeting & fix time though discussion. I need to observe either all members participated in the meetings. I also think about how community clinics should run and maintenance of drug in the community clinics. I provide a box for raising fund in our community clinic & by one to two taka makes so much to buy a chair and a table and If there any treatment of poor patient*.*”*

During FGD another CSG member said *“At first we search pregnant mothers & we made courtyard meeting with them, their husband, mother in laws and mother and motivate as many responsibilities & preparation to be a father, mother or grandmother & advice them to provide extra food and regular health checkup to the mother*.*”*

During IDI one of the CSG shares *“I collect money for support of someone have deficiency of money. If there any problem occurs then there is no need for meeting, in need everyone worked as team to help people. I told them for antenatal checkup four times during pregnancy and after delivery. I feel so pain when I saw a disabled boy near my house and his mother always busy for take care of him. This is because the mother was not conscious about the checkup during pregnancy*.*”*

During IDI one CG member said *“First duty is to keep this clinic for long lasting, to solve the problem of the clinic. We made a meeting for salvation of electricity & toilet problem; I gave advice to come clinics for vaccination, collecting fund, to aware about pregnancy problem and clinics facilities. I also help during road construction by giving worker for 10 days working. We conduct meeting at every one or two months interval at the house of pregnant woman where we advice them about regular checkup & avoid risky works during pregnancy*.*”*

#### Barrier on the duties as CG/CSG

In response about the obstacle faced during duties and responsibilities of CG and CSG, most of them expressed various problem but few never face any obstacle. The community people avoid them when they said about family planning and wanted to know about our salary and also asked about their financial benefit. Maximum old women ignore the advices related to maternal & neonatal care as they believed there is no problem occurred during pregnancy even with hard work. Husbands of pregnant mothers ignore to take their wife at facility and emphasis for traditional delivery at home.

During IDI one CSG member shares *“Husband and mother in law of the pregnant mothers never want to listen. Sometime mother in laws said that she had no problem during her delivery of even seven children with hard work during pregnancy. Some husband said that there is no need to go to doctor and delivery will occur traditionally in home. When we said to the mother in laws about to call us if there any emergency then rashly answered as what we can do for them and said that they can solve problem by themselves. They always seek financial benefit in spite of taking advice”*

During IDI one CG member mentioned “*No, there has been problem as I provide drug to them they come to me for advice but rich people go to Sathkhira & Kulaora hospital and never come to us for any support”*

During FGD one CSG member said *“nobody makes any trouble. I work for them without my personal benefit, such as I asked school head master for giving a chair. If I face any problem then I call volunteer to make a meeting and solving the problem, all of us get together and solve the problem. But I feel so bad sometime that for whom we are doing a lot they are doing nothing for us, not even give us ten taka*.*”*

#### Practices to overcome the obstacle

In response to how the CG/CGS overcome these obstacle most of them said that they solved the obstacles through discussion with others members and higher authority and also by convincing the community people by various way.

During IDI One CG member said *“our meeting can make a solution, if not possible then we inform to the person who are available at community clinics and get solutions. We didn’t give any chance to arise a problem. Though we have no operation facility at community clinic we manage the vehicle for referral of emergency patients*.*”*

During FGD one CSG member said *“The main problem is that the community people didn’t give importance when we went at first time, but we have to go every house for 5 to 7 times and then they asked me about the objectives and listen to me*.*”*

During FGD another CSG member said *“sometime we call a meeting and collect 200-250 taka from which 50 taka given to the fund for savings and solving many problems. I solved the problem through continuously appreciate the community people of their activities*.*”*

##### How to improve the mother and child’s health

In response of what else can do for the improvement of mother and child’s health most of CG and CSG member said about the necessity of a gynecology doctor, availability of essential drugs, transport facilities and improvement of delivery room at their community clinic. Few of them raise the issues of separate funding system and trained up the staffs and CG/CSG member.

During IDI one CG member said *“If there is a qualified doctor in this Community clinic then we can collect 100 taka from each patient that can be utilized for wall and gate construction and payment of night guard. 24 hours electricity should ensure here and skilled person need for delivery with all equipment. The road should reconstruct for referral of emergency patient*.*”*

During FGD one of CG member said *“there should be a qualified doctor to handle the critical patient at community clinic. A tube well and toilet is essential here. There should be improve the delivery facilities, a multistoried building and as Kularoa is far away so number of bed and drug should increases*.*”*

During FGD one CSG member said *“there is necessary of a delivery specialist in our community clinic with separate room of delivery. Electricity and vehicle should available and if a MBBS doctor is appointed here then we don’t need to go at Kularoa*.*”*

During IDI one CSG member said *“In our community clinic both the facility of normal delivery or caesarean operation should available. We need the facility of emergency oxygen and blood*. We *should aware first, then we can aware others. The clinic should be secured by fencing and if a MBBS doctor is available then the mothers need not to go far for emergency treatment*.*”*

## Discussion

The findings from the Focus Group Discussions (FGDs) conducted in Kularoa upazila of Sathkhira district shed light on the disparities in knowledge and practices regarding maternal and neonatal health (MNH) services between mothers in intervention and control areas. This discussion aims to analyze these findings and highlight the significance of community-based interventions in improving MNH outcomes.

Mothers in the intervention area exhibited higher levels of knowledge and proactive practices regarding birth planning, pregnancy complications, and antenatal care (ANC)^11^. They engaged in birth planning, received ANC regularly, and were aware of maternal complications. In contrast, mothers in the control area lacked awareness of birth planning and maternal complications, often perceiving them as normal occurrences during pregnancy. They relied on traditional birth attendants for delivery. Another, grave situation for adolescent female in Bangladesh is early marriage and pregnancy which in term leads malnutrition of both child and mother. The ultimate consequence would be increased maternal and child mortality^12^. It is in the top ten nations in terms of both absolute and relative pregnancy rates among adolescent females. A study showed that adolescent mothers had worse maternal nutritional status and took longer to recover after childbirth despite getting the same amount of maternal health and nutrition interventions as adult pregnant women^13^.

Intervention area mothers actively planned for delivery, including selecting the place and person for delivery, receiving ANC, and preparing for emergencies^14,15^. Control area mothers, however, showed minimal emphasis on birth planning, often opting for home deliveries attended by traditional birth attendants^16^.

Mothers in the intervention area received ANC regularly, including health checkups and tests^17^. They were aware of pregnancy complications and sought medical help when necessary^18^. Control area mothers lacked clear ideas about ANC and only sought medical assistance in case of complications, often relying on traditional beliefs^19^.

Intervention area mothers benefited from community support groups and volunteers who provided essential information and assistance during pregnancy and childbirth. Control area mothers had limited knowledge of community support structures and lacked engagement with volunteers, leading to missed opportunities for health education and support.

Both community volunteers and support group members faced obstacles such as resistance from community members, especially regarding traditional beliefs and financial expectations. Overcoming these challenges required continuous engagement, education, and collaboration with community leaders and stakeholders. According to a UNICEF Bangladesh Situation Analysis from 2015, there are numerous interconnected and intersecting determinants that prevent children and adolescents from surviving, developing, being protected, and participating in society across all age groups and sectoral areas. This is especially true from the perspective of behaviors and social change. Inadequacies in the structural or enabling environment to have been noted regarding traditional social norms, gender differences and oppressive social dynamics between the sexes, insufficient laws, policies, and development strategies and resources, among others^20^.

Enhancing the availability and quality of MNH services at community clinics, including 24-hour services, skilled personnel, essential drugs, and transportation facilities. Providing incentives or honorariums for community volunteers and support group members to enhance their motivation and commitment. Conducting regular training and capacity-building programs for volunteers and support group members to improve their effectiveness in delivering MNH services.

## Conclusion

The findings underscore the importance of community-based interventions in promoting maternal and neonatal health. By addressing knowledge gaps, improving access to essential services, and fostering community engagement, these interventions have the potential to significantly reduce maternal and neonatal morbidity and mortality rates. Continued investment in such initiatives is essential for achieving sustainable improvements in MNH outcomes in resource-constrained settings like Kularoa upazila.

## Data Availability

All the data and voice recordings are available based on request type.

## Notes

### Competing Interest Statement

The authors have declared no competing interest.

### Author Declarations

Ethical approval was obtained from the Ethical Review Committee (ERC) of the Centre for Injury Prevention and Research, Bangladesh (CIPRB) before commencement of the study (CIPRB/ERC/2020/11, Date: 29 January 2020).

